# Stratified assessment for geriatric mental health using probabilistic graphical model: a cross-sectional observational study in India

**DOI:** 10.1101/2023.03.26.23287749

**Authors:** Indra Prakash Jha, Shreya Mishra, Neetesh Pandey, Vibhor Kumar

## Abstract

**Background:** The mental health among seniors presents a diverse set of challenges that can be attributed to a number of different causes, including social factors, economic factors, and psychological factors. Researchers are attempting to understand its complexity; however, there is a limited amount of systematic robust analysis that has been conducted. It is, therefore, critical to identify and quantify the driver characteristics or factors that are related to mental health and conduct age-specific stratified analysis in order to guide and provide tailor-made policy-level interventions.

**Methods:** We used “Longitudinal Ageing Study in India (LASI) wave-1 survey” data of 72250 (47% male and 53% female) carried out in April 2017 to December 2018 spanning over 35 states and union territories with socioeconomic, behavioural, psychological, and health-related indicators. We utilised a probabilistic graphical model named Bayesian Network to identify key factors of mental health variables.

**Results:** Ageism, social discrimination (the inferred conditional probability difference is ∼30% with 95% CI: ∼5%) and Food insecurity (>19% with a 95% CI of >3%) were found to negatively impact mental health in older adults, while connectedness with family and friends (9.34% with 95% CI: 1.41%) positively affected mental health. The sense of general dissatisfaction or discontentment with one’s own life (17% with 95% of CI: ∼2.8%) is one of the factors that plays a significant role in mental health among elderly people. Additionally, multimorbidities of mental health problems in older adults, which were found to be arthritis, sleep disorders, and stroke. The incidence of multimorbidity due to mental issues were shown to be higher in older adults than younger adults, with sleep disorders being associated with mental health in older age groups. By incorporating age based stratification analysis, we also identified the specific age group when older people from India are most vulnerable to mental health issues.

**Conclusion:** The prevalence of mental illness in senior citizens in India needs to be reduced, and one way to do this is through the development of targeted policies to combat food insecurity and social discrimination, as well as to improve communication of older people with their loved ones. In addition to this, the issue of arthritis must be tackled in a more in-depth manner given the result that it also has a severe impact on the mental health among elderly.

## Introduction

Mental wellbeing is a critical aspect of healthy aging. The elderly population is particularly vulnerable to mental health issues. As the world’s population aging rapidly, it is crucial to understand the prevalence as well as the drivers of seniors’ mental health issues. Studies have shown that mental health problems can have a substantial impact on one’s daily routine lifestyle, functional status, and overall health outcomes in this population [1,2]. The global population of older adults is expected to increase rapidly in the coming decades, with a projected increase from 703 million people having age 65 or above in 2019 to 1.5 billion in 2050 [3]. India is projected to have the second largest population of older adults in globe, with an estimated 340 million people having age 65 or above by 2050 [4] [5]. With this rise in the number of elderly people, it is anticipated that the burden of mental health problems among older persons will also increase, posing a huge public health challenge in India. This rapid ageing has implications for global health and healthcare systems, as the elderly population is more susceptible to chronic diseases, disabilities, and mental health problems [6,7].

Research has shown that mental health issues are prevalent among the elderly population worldwide. Research [8] discovered that the prevalence of depression among senior citizens was 7.2% in low-income and middle-income countries, and 11.5% in high-income countries. These findings highlight the need for research on mental health issues among the elderly population, specially in low- and middle-income countries where the elderly population is rapidly growing. Society must pay greater consideration to the various mental and physical health issues faced by the elderly. Nearly 20% of older people, having age 60 or above, have some sort of mental or neurological disorder (excluding headache disorders) [9,10]. Mental and neurological disorders accounted for 6.6% of all DALYs (disability-adjusted life years) among people aged 60 and above [11], and according to recent reports, mental, neurological, and substance abuse problems constituted for 10.5% of the global disease challenge, as measured by DALYs [12–14]. The YLDs for mental and substance abuse problems increased by 45% from 1990 to 2013 [14]. Dementia and depression are the most prevalent mental and neurological illnesses among the elderly. Around 5% and 7%, respectively, of the world’s elderly people suffer from dementia or depression. Due to heavy social and economic problems, these problems are worse in developing countries like India [15,16]. Mental health problems aren’t always noticed by doctors or by older people themselves, and the stigma that comes with them makes people less likely to ask for help.

Additionally, bereavement and retirement-related declines in socioeconomic status appear to be more common in older people [17]. Each of these stress factors has the potential to make older people feel isolated, lonely, or distressed psychologically, necessitating long-term care. Physical and psychological health are related [18,19]. Studies in the past have shown that there is a significant relationship between socioeconomic status (SES) and mental health in many ways. Seniors in countries with high income inequality, for example, have worse mental health than seniors in countries with low income inequality. Adults living in urban areas were also found to be negatively impacted by displacement, demonstrating the link between SES and mental health. As socioeconomic status (SES) improved, the incidence of poor mental health among the elderly in Japan reduced, suggesting a favourable relationship among SES and mental wellbeing. The psychological resources of older people, such as their sense of autonomy, self-efficacy, and optimistic self-beliefs, are impacted by their socioeconomic status (SES) [20]. For example, self-efficacy [21] might motivate older persons to practise mentally healthy lifestyles because they feel capable of managing the expectations and obstacles of their surroundings. In a way analogous, elderly people who are optimistic about the future regularly encounter a healthier lifestyle. Furthermore, it is believed that older adults’ sense of independence is beneficial to their health because it lessens the harmful impact of stress factors. There has not been much research that looks at how much the socioeconomic status of a neighbourhood influences self-efficacy, which in turn influences health. The vast majority of studies that have been conducted in this area have focused only on examining how self-efficacy [22] influences the association between SES variables and health on an individual level. Social instruments like social protection, trust between people, and solidarity can protect elderly individuals with mental illnesses from the harmful consequences of discrimination. For example, if people had access to instrumental (like help with travel), informational (like information about a health problem), or affective (like comforting in the face of a health problem) social support, they might be able to get some resources that they wouldn’t be able to get otherwise. These examples that social support keeps people safe also show how important it is to look at the surroundings. Social environments like social solidarity in a neighbourhood are especially important sources of social support.

While there has been substantial research into the correlation between socioeconomic status and psychological well-being, there has been surprisingly little investigation into the effects of demographic diversity in these models. In the current work, we analysed information collected from an extensive survey of the Indian elderly population, covering areas such as language, culture, and other dimensions of diversity. Using these, we first used bayesian network inferences to quantify the relationship of socioeconomic status (SES) and mental wellbeing. Furthermore, we utilised the same method to evaluate the comorbidities of mental health with other chronic diseases as well. As far as we know, this is the first time Bayesian networks, a type of graphical model, have been used to look into the mental health of India’s older people. Our research has the potential to guide targeted interventions and policies to promote healthy aging and improve mental health outcomes in this population by identifying the factors associated with these outcomes. Because of the rapid increase in the world’s elderly population, our findings may also be relevant to other low and middle-income countries where these problems are rampant.

To further investigate, we also performed the Age-based stratification analysis. The Age-based stratification analysis is important for the society because it helps to identify the specific needs and vulnerabilities of different age groups, particularly the elderly population, who are more vulnerable to multimorbidity of mental disorder. By gaining a deeper understanding of the patterns and prevalence of mental illnesses among older adults, the policymakers, healthcare providers, and social service agencies can better allocate resources and design interventions that target the specific needs of this population.

Age-based stratification analysis also helps to identify possible risk attributes and preventive attributes related to older adults’ mental health. This information can inform the development of preventive measures and early interventions to reduce the prevalence and severity of older adults’ mental health. Moreover, understanding the impact of multimorbidity of mental health on older adults can inform the design of effective and equitable health and social care policies that consider the unique needs of this population.

In short, age-based stratification analysis is important for society because it provides valuable insights into the mental health needs of older adults, which can inform the design of targeted interventions, policies, and programs to enhance their overall health and life as a whole.

The rest of this paper is structured as follows: first, we understand the overview of the LASI dataset and the methods used in our study. After that, we show the results of our analysis and talk about what they mean. Lastly, we talk about the problems with our study and ways to do more research in the future.

## Methods

### Study Population

The data came from the Longitudinal Ageing Study in India (LASI) Wave-1 survey [23] [24], which included samples of 72,250 individuals having age 45 or above and their partners, including 31,464 elderly (age 60 or above) and 6,749 oldest-old people (age 75 or above) from India. The data is distributed from thirty five states and union territories (except Sikkim) of the country. The said survey was conducted throughout the country from April 2017 to December 2018. The LASI is a “multistage stratified area probability cluster sampling design” to find the eventual observations: older adults aged 45 or above along with partner (without considering their partner’s age). The original data’s specifics can be found elsewhere [25]. Table 1 shows the social, economic, and demographic information about the people who took part in the survey.

**Table 1:** Demographic and socio-economic characteristics.

| variables | Male | Female | Overall |
| --- | --- | --- | --- |
| **State** |  |  |  |
| (Row %)(Col %) | n = 30569 (42.31%) | n = 41681 (57.69%) | n = 72250 (100%) |
| Andaman & Nicobar Islands | 566 (45.50%) (1.85%) | 678(54.50%)(1.63%) | 1244(100%)(1.72%) |
| Andhra Pradesh | 1120 (41.81%) (3.66%) | 1559(58.19%)(3.7%) | 2679(100%)(3.71%) |
| Arunachal Pradesh | 552 (45.43%) (1.81%) | 663(54.57%)(1.59%) | 1215(100%)(1.68%) |
| Assam | 973 (41.12%) (3.18%) | 1393(58.88%)(3.3%) | 2366(100%)(3.27%) |
| Bihar | 1555 (44.18%) (5.09%) | 1965 (55.82%) (4.71%) | 3520 (100%) (4.87%) |
| Chandigarh | 452 (44.05%) (1.48%) | 574 (55.95%) (1.38%) | 1026 (100%) (1.42%) |
| Chhattisgarh | 906 (44.09%) (2.96%) | 1149 (55.91%) (2.76%) | 2055 (100%) (2.84%) |
| Dadra & Nagar Haveli | 475 (43.58%) (1.55%) | 615 (56.42%) (1.48%) | 1090 (100%) (1.51%) |
| Daman & Diu | 410 (41.37%) (1.34%) | 581 (58.63%) (1.39%) | 991 (100%) (1.37%) |
| Delhi | 599 (45.41%) (1.96%) | 720 (54.59%) (1.73%) | 1319 (100%) (1.83%) |
| Goa | 568 (39.80%) (1.86%) | 859 (60.20%) (2.06%) | 1427 (100%) (1.98%) |
| Gujarat | 989 (42.25%) (3.24%) | 1352 (57.75%) (3.24%) | 2341 (100%) (3.24%) |
| Haryana | 792 (41.73%) (2.59%) | 1106 (58.27%) (2.65%) | 1898 (100%) (2.63%) |
| Himachal Pradesh | 562 (40.49%) (1.84%) | 826 (59.51%) (1.98%) | 1388 (100%) (1.92%) |
| Jammu & Kashmir | 711 (44.08%) (2.33%) | 902 (55.92%) (2.16%) | 1613 (100%) (2.23%) |
| Jharkhand | 1043 (42.33%) (3.41%) | 1421 (57.67%) (3.41%) | 2464 (100%) (3.41%) |
| Karnataka | 973 (40.21%) (3.18%) | 1447 (59.79%) (3.47%) | 2420 (100%) (3.35%) |
| Kerala | 991 (39.69%) (3.24%) | 1506 (60.31%) (3.61%) | 2497 (100%) (3.46%) |
| Lakshadweep | 446 (39.16%) (1.46%) | 693 (60.84%) (1.66%) | 1139 (100%) (1.58%) |
| Madhya Pradesh | 1324 (45.44%) (4.33%) | 1590 (54.56%) (3.81%) | 2914 (100%) (4.03%) |
| Maharashtra | 1607 (40.45%) (5.26%) | 2366 (59.55%) (5.68%) | 3973 (100%) (5.50%) |
| Manipur | 581 (42.44%) (1.90%) | 788 (57.56%) (1.89%) | 1369 (100%) (1.89%) |
| Meghalaya | 368 (37.98%) (1.20%) | 601 (62.02%) (1.44%) | 969 (100%) (1.34%) |
| Mizoram | 563 (45.18%) (1.84%) | 683 (54.82%) (1.64%) | 1246 (100%) (1.72%) |
| Nagaland | 589 (44.76%) (1.93%) | 727 (55.24%) (1.74%) | 1316 (100%) (1.82%) |
| Odisha | 1255 (43.02%) (4.11%) | 1662 (56.98%) (3.99%) | 2917 (100%) (4.04%) |
| Puducherry | 552 (38.66%) (1.81%) | 876 (61.34%) (2.10%) | 1428 (100%) (1.98%) |
| Punjab | 927 (43.64%) (3.03%) | 1197 (56.36%) (2.87%) | 2124 (100%) (2.94%) |
| Rajasthan | 970 (43.23%) (3.17%) | 1274 (56.77%) (3.06%) | 2244 (100%) (3.11%) |
| Tamil Nadu | 1394 (39.49%) (4.56%) | 2136 (60.51%) (5.12%) | 3530 (100%) (4.89%) |
| Telangana | 1008 (40.73%) (3.30%) | 1467 (59.27%) (3.52%) | 2475 (100%) (3.43%) |
| Tripura | 491 (41.09%) (1.61%) | 704 (58.91%) (1.69%) | 1195 (100%) (1.65%) |
| Uttar Pradesh | 2081 (45.57%) (6.81%) | 2486 (54.43%) (5.96%) | 4567 (100%) (6.32%) |
| Uttarakhand | 556 (40.94%) (1.82%) | 802 (59.06%) (1.92%) | 1358 (100%) (1.88%) |
| West Bengal | 1620 (41.19%) (5.30%) | 2313 (58.81%) (5.55%) | 3933 (100%) (5.44%) |
| **Residence i.e Rural/Urban ?** |  |  |  |
| (Row %)(Col %) | n = 30569 (42.31%) | n = 41681 (57.69%) | n = 72250 (100%) |
| Rural | 19945 (42.86%) (65.25%) | 26589 (57.14%) (63.79%) | 46534 (100%) (64.41%) |
| Urban | 10624 (41.31%) (34.75%) | 15092 (58.69%)<br>(36.21%) | 25716 (100%) (35.59%) |
| **Living arrangements i.e. Living with ?** |  |  |  |
| (Row %)(Col %) | n = 30569 (42.31%) | n = 41681 (57.69%) | n = 72250 (100%) |
| living alone | 591 (25.55%) ( 1.93%) | 1722 (74.45%) (4.13%) | 2313 (100%) ( 3.20%) |
| living with spouse and/or others | 5429 (50.09%) (17.76%) | 5409 (49.9%) (12.98%) | 10838 (100%) (15.0%) |
| living with spouse and children | 21007 (48.1%) (68.72%) | 22656 (51.8%)(54.36%) | 43663 (100%) (60.43%) |
| living with children and others | 2536 (20.30%) ( 8.30%) | 9958 (79.7%) (23.89%) | 12494 (100%) (17.29%) |
| living with others only | 1006 (34.19%) ( 3.29%) | 1936 (65.81%) ( 4.64%) | 2942 (100%) ( 4.07%) |
| **MPSE Quintile ie. Economic Well Being** |  |  |  |
| (Row %)(Col %) | n = 30569 (42.31%) | n = 41681 (57.69%) | n = 72250 (100%) |
| Poorest | 5976 (42.21%) (19.55%) | 8182 (57.7%)(19.63%) | 14158 (100%) (19.60%) |
| Poorer | 6103 (42.00%) (19.96%) | 8427 (58.00%) (20.22%) | 14530 (100%) (20.11%) |
| Middle | 6119 (42.09%) (20.02%) | 8418 (57.91%) (20.20%) | 14537 (100%) (20.12%) |
| Richer | 6227 (42.40%) (20.37%) | 8459 (57.60%) (20.29%) | 14686 (100%) (20.33%) |
| Richest | 6144 (42.85%) (20.10%) | 8195 (57.15%) (19.66%) | 14339 (100%) (19.85%) |
| **Education** |  |  |  |
| (Row %)(Col %) | n = 30569 (42.31%) | n = 41681 (57.69%) | n = 72250 (100%) |
| Less than Primary (Standard 1-4) | 4069 (50.52%) (13.31%) | 3986 (49.48%) ( 9.56%) | 8055 (100%) (11.15%) |
| Primary Completed (Standard 5-7) | 4738 (48.94%) (15.50%) | 4943 (51.1%) (11.86%) | 9681 (100%) (13.40%) |
| Middle Completed (Standard 8- 9) | 3874 (53.60%) (12.67%) | 3353 (46.40%) ( 8.04%) | 7227 (100%) (10.00%) |
| Secondary School/Matriculation completed | 3849 (57.26%) (12.59%) | 2873 (42.74%) ( 6.89%) | 6722 (100%) ( 9.30%) |
| Higher Secondary/Intermediate/Senior Secondary completed | 1925 (59.34%) ( 6.30%) | 1319 (40.66%) ( 3.16%) | 3244 (100%) ( 4.49%) |
| Diploma and certificate holders | 244 (71.55%) ( 0.80%) | 97 (28.45%) ( 0.23%) | 341 (100%) ( 0.47%) |
| Graduate degree (BA., BSc., BCom) awarded | 1540 (62.10%) ( 5.04%) | 940 (37.90%) ( 2.26%) | 2480 (100%) ( 3.43%) |
| Post-graduate degree or (MA., MSc., M. Com.) higher (MPhil, PhD, PostDoc) awarded | 471 (62.72%) ( 1.54%) | 280 (37.28%) ( 0.67%) | 751 (100%) ( 1.04%) |
| Professional course/degree (BEd, B.Tech, MBBS, BHMS, BAMS, BPharm, BCA, BBA, LLB, MTech, MD etc)awarded | 363 (67.47%) ( 1.19%) | 175 (32.53%) ( 0.42%) | 538 (100%) ( 0.74%) |
| Never Attended The School | 9495 (28.59%) (31.06%) | 23712 (71.4%) (56.8%) | 33207 (100%) (45.96%) |
| Not Provided | 1 (25.00%) ( 0.00%) | 3 (75.00%) ( 0.01%) | 4 (100%) ( 0.01%) |
| **Religion** |  |  |  |
| (Row %)(Col %) | n = 30569 (42.31%) | n = 41681 (57.69%) | n = 72250 (100%) |
| Hindu | 22483 (42.4%) (73.5%) | 30490 (57.5%) (73.1%) | 52973 (100%) (73.3%) |
| Muslim | 3555 (41.02%) (11.63%) | 5112 (58.9%) (12.26%) | 8667 (100%) (12.00%) |
| Christian | 3041 (42.15%) ( 9.95%) | 4174 (57.8%) (10.01%) | 7215 (100%) ( 9.99%) |
| Sikh | 883 (44.17%) ( 2.89%) | 1116 (55.83%) ( 2.68%) | 1999 (100%) ( 2.77%) |
| Buddhist/neo-Buddhist | 224 (42.34%) ( 0.73%) | 305 (57.66%) ( 0.73%) | 529 (100%) ( 0.73%) |
| Jain | 77 (43.75%) ( 0.25%) | 99 (56.25%) ( 0.24%) | 176 (100%) ( 0.24%) |
| Jewish | 2 (40.00%) ( 0.01%) | 3 (60.00%) ( 0.01%) | 5 (100%) ( 0.01%) |
| Parsi/Zoroastrian | 4 (57.14%) ( 0.01%) | 3 (42.86%) ( 0.01%) | 7 (100%) ( 0.01%) |
| Religion other than above Other | 233 (44.05%) ( 0.76%) | 296 (55.95%) ( 0.71%) | 529 (100%) ( 0.73%) |
| No Religion | 66 (45.52%) ( 0.22%) | 79 (54.48%) ( 0.19%) | 145 (100%) ( 0.20%) |
| Not Provided | 1 (20.00%) ( 0.00%) | 4 (80.00%) ( 0.01%) | 5 (100%) ( 0.01%) |

### Analysis

#### Assessment of mental health

For the analysis, two different estimates were used for mental health. One, for the purpose of multimorbidity analysis among diagnosed chronic diseases for mental health, the attribute ‘HT009’ is used, which refers to “any neurological or psychiatric problems such as depression, Alzheimer’s or dementia, unipolar or bipolar disorders, convulsions, Parkinson’s, etc. ever diagnosed by a health professional”. Second, the Centre for Epidemiologic Studies Depression (CES-D) scale15 was used to determine the existence of psychological distress in order to conduct further research into the associations of socioeconomic factors, discrimination, and family network structure with elderly mental health (Radloff, 1977). CESD is a “short self-reported scale designed” as a general population based screening instrument for depressive symptoms [26].

#### Multimorbidity Network analysis

The occurrence of two or more chronic illnesses simultaneously is referred to as “multimorbidity” by medical practitioners. It is common knowledge that the risk of having multiple diseases increases with age. People who have more than one disease tend to lose their physical and functional abilities more quickly and have a higher chance of dying. In this study, we used Bayesian Network Analysis [27] [28] [29] to analyse the multimorbidity pattern of elderly people graphically. We included diseases such as cardiovascular-diseases CVD (including one or more from hypertension HT or high blood pressure, stroke, and chronic heart diseases), diabetes mellitus, high cholesterol, chronic lung diseases (including one or more form asthma, chronic obstructive pulmonary disease COPD, Chronic bronchitis), bone/joint diseases (include one or more from Arthritis, rheumatism, Osteoporosis), cancer or malignant tumour, Thyroid disorder, Gastrointestinal problems (including one or more of GERD, constipation, indigestion, piles, andeptic ulcers), skin disorders, urogenital problems (include one or more from Chronic Renal incontinence continence, Kidney Stones, Benign Prostatic Hyperplasia BPH), sleep disorder. And, then learned the joint probabilistic association using a bayesian network with mental disorders to assess multimorbidity graphically.

#### Association of Socioeconomic status Factors with Mental-Health

The analysis of socioeconomic status (SES) factors in relation to mental health has revealed important associations. This relationship has been found to be particularly strong among older adults. Many factors, including financial stress, social isolation, and fewer opportunities, are believed to contribute to the association of SES and mental health. When creating interventions to support mental wellbeing and lower the challenge of mental illness, it is vital to incorporate SES characteristics, particularly among disadvantaged populations. Data had a spectrum of SES and demographic attributres: age, gender, marital status (married, unmarried), education level, languages and work, retirement, and pension status; Chronic Health conditions: general health (“Excellent / Very good/good” or “fair/poor”), chronic conditions (cardiovascular diseases, chronic heart diseases, stroke, cancer, urogenital, etc.) other than mental health; Functional Health: Limitations and helpers (physical and psychological impairment, mobility, activities of daily living ADL, instrumental activities of daily living IADL, support from devices); family and social network (living arrangements, social and family connectedness, social support); decision-making, social participation, and life satisfaction (ageism/discrimination, life satisfaction). The primary outcome of interest was to learn about the impact of socioeconomic factors based on different levels of mental health (rarely or never, sometimes, often, most, or all of the time) for variously identified covariates. Using Bayesian network inferences, we used socioeconomic indicators from the data to measure the relationship between these indicators and mental health.

#### Age based stratification analysis for SES and Multimorbidity

After gaining an understanding of multimorbidity as well as the association of socioeconomic status and mental health, we carried out an age-based stratification analysis. Our research shows that both the effect of socioeconomic status (SES) on mental health and the number of co-morbidities related to mental health change with age. The same socioeconomic factors can have very different effects on people of different ages, with some age groups being more susceptible to mental health problems and multimorbidity than others. For instance, elderly people between the ages of 45 and 64 have the lowest risk of being exploited due to a lack of social support. In light of this, it is essential to take into account age-based stratification when investigating the connection between socioeconomic status, mental health, and the presence of multiple diseases. When these associations are understood, targeted interventions and policies can be developed to lessen the impact of mental health problems and multiple diseases on people of varying ages.

## Results

### Exploratory Analysis

We systematically analyzed the survey of 72250 (42% male and 57% female) elderly adults. It has been found that the prevalence of mental health is almost identical in its distribution, gender-wise, economic wellbeing condition-wise (MPSE_Quintile), or living place-wise (whether rural or urban).

### Multimorbidity Network analysis: Chronic Diseases and Mental Health

From the Bayesian network inference, we discovered that mental health problems are directly comorbid with arthritis, sleep disorders, and stroke. The conditional probability difference, by applying exact inference, for arthritis it is ∼20% (with 95% CI:∼5%), for sleep disorders it is ∼18% (with 95% CI:∼5%), for stroke it is >6%(with 95% CI:∼3%) and for hypertension it is >5.5%(with 95% CI:∼5%).

#### Age based stratification analysis: Multimorbidity

Age-based stratification refers to the differential distribution of multimorbidity across different age groups. In the context of mental health, age-based stratification has important implications for healthcare planning and delivery. We discovered that the prevalence of mental health multimorbidity is significantly higher among older adults in comparison to younger adults. For individuals with ages less than or equal to 45, the Bayesian network suggests that their mental health is associated with arthritis and stroke. On the other hand, the Bayesian network for individuals with ages ranging from 45 to 64, 65 to 75, and over 75 suggests that their mental health is associated not only with arthritis and stroke but also with sleep disorders [Figure 2]. As a result, it is possible to conclude that ageing contributes to the additional association between sleep disorder and mental health.

**Figure 1:**
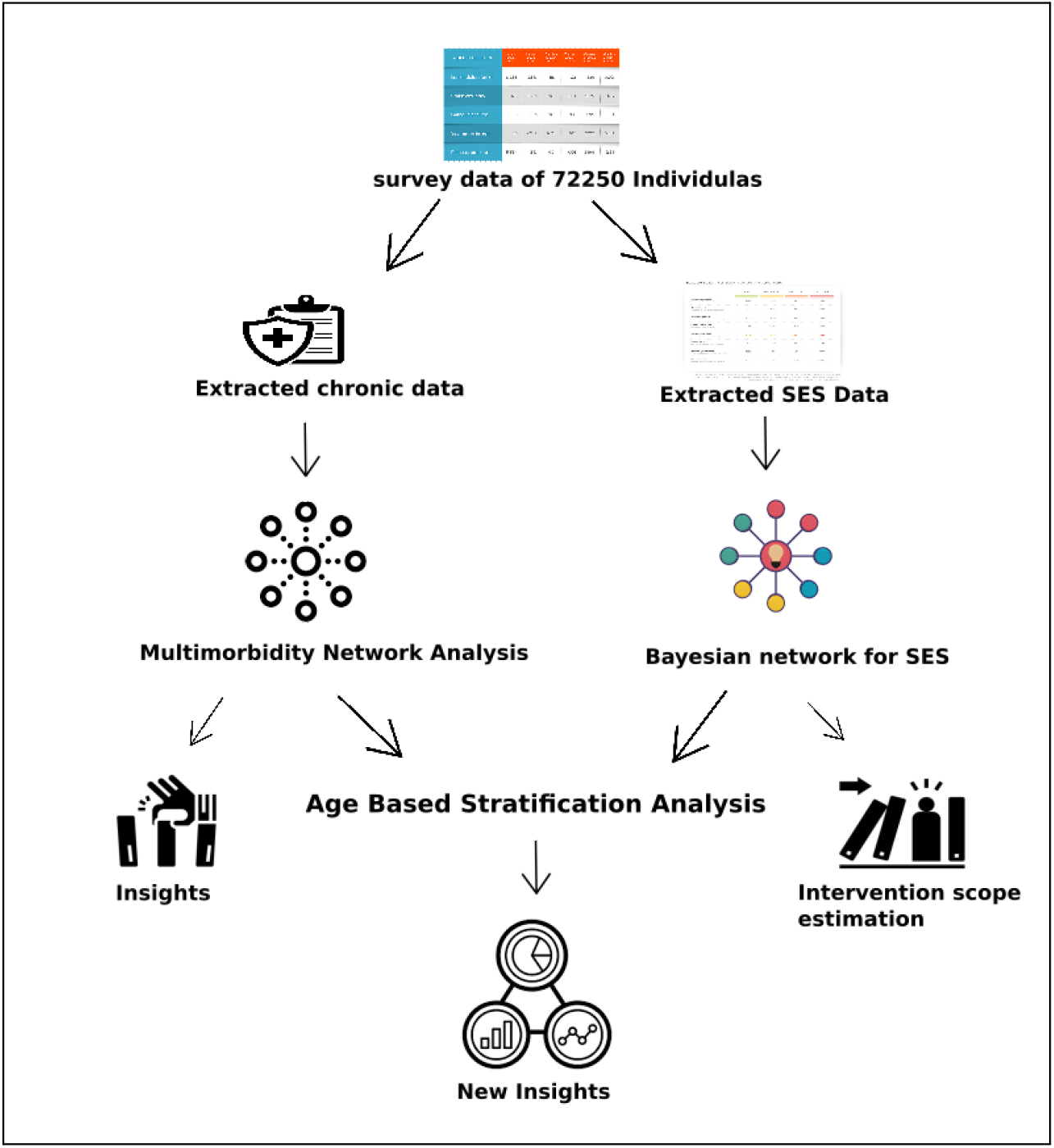
The pipeline of our methodological approach. Using a feature-wise partition, we separated data from 72250 individuals into two groups. First, socioeconomic (SES) and demographic variables are retrieved, then chronic illness variables. The Bayesian network analysis was utilised to analyse the first dataset for multimorbidity and to generate insights into multimorbid chronic mental health disorders. Subsequently, again utilising Bayesian network analysis, we searched the second partitioned dataset for socioeconomic drivers of psychological health. These clarifying characteristics may aid in determining the intervention’s scope. Then, we conducted an additional study that shed insight on the subject at hand: an age-based stratification analysis.

**Figure 2:**
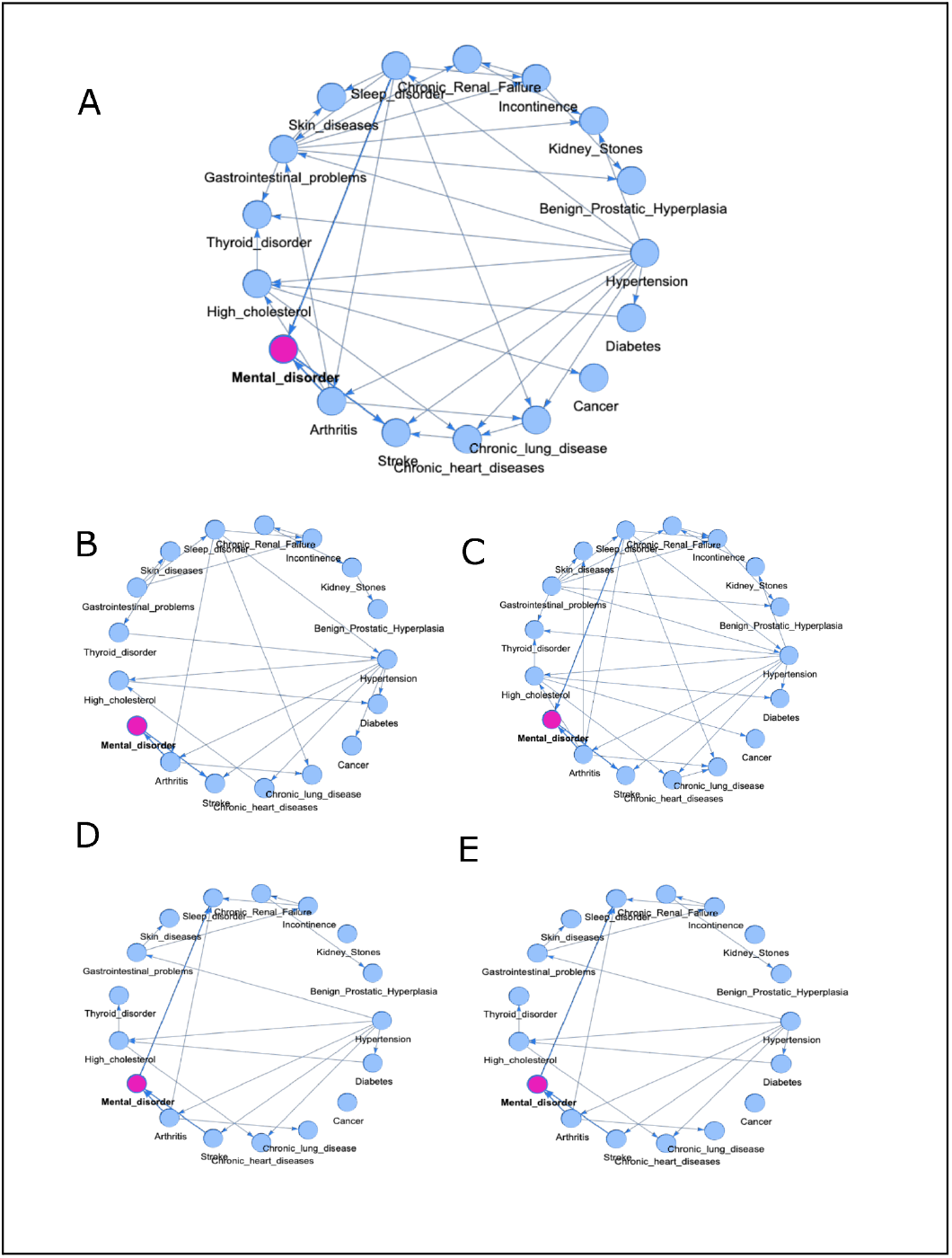
Multimorbidity Analysis: (A) A Bayesian network of all 17 diseases together, consisting of the complete dataset. The goal was to find out which chronic diseases are also linked to mental health problems. (B) Bayesian network for individuals having ages less than or equal to 45. It is evident that mental health is not associated with sleeping disorders here. (C) Bayesian network for people between the ages of 45 and 64. Here, one additional linkage associated with mental health is found with sleep disorders. (D) Bayesian network for people between the ages of 65 and 74. Here, one additional linkage associated with mental health is found with sleep disorders. (E) A Bayesian network for people over the age of 75. Here also one additional linkage associated with mental health is found with sleep disorders.

### Data-Driven Bayesian Network Analysis

#### Social Factor (Social Discrimination)

Ageism is a widespread and alarming problem in the world today. It affects both developed and developing countries. Multiple instances of social discrimination are reported in day-to-day life by older people. The associations with the seniors’ mental health can be seen as differential in inferred conditional probability. Older people are facing social discrimination in various different forms, such as receiving less courtesy or respectful treatment than other people (∼30% with 95% CI: ∼5%), poorer service than other people at public places like restaurants or stores (∼29% with 95% CI: 6.5%), being threatened or harassed (30.8% with 95% CI: 7.06%), and even being treated as if they are not smart (∼34% with 95% CI: 5.27%).

#### Economical Factor (Food Insecurity)

Our analysis showed a clear impact of food security on an individual’s mental health. Food security has been linked to depression in a variety of ways, including subjects having to reduce their meal size due to a lack of food (>19% with a 95% CI of >3%) or being hungry but not eating due to a lack of food (4.8% with a 95% CI of 1.8%). Even so, it has also been found that because of hunger and a lack of food, the subject lost weight, which, in turn, affected their mental health (3.68% with a 95% CI: 1.3%).

#### Psychological Factor (Discontentment of Life)

A widespread sense of discontent with one’s life is one of the factors that plays a significant role in the mental health of the elderly population. The conditional probability of depression is 17% [95% of CI: ∼2.8%] higher among older people who have experienced general dissatisfaction with life (95% confidence interval: 2.8% lower).

#### Family and friends connectedness Factor (Communication)

Connectedness with family and friends is also found to be an important factor that positively affects the mental health of older people. The differential inferred conditional probability for older people who regularly meet with friends was 4.5% [95% CI: 0.4%], whereas for people who communicate and share personal matters with their partner/spouse, differential inferred conditional probability is 9.34% [95% CI: 1.41%]. It is also worth noting that virtual communication using phone, email, etc. has less effect on mental health but also improves it [3.4% with a 95% CI: 0.2%].

#### Age based stratification analysis: Association of SES with mental health

We also performed an age-based stratification analysis to learn more about the association between different factors and mental health [Figure 3]. We found that social factors (social discrimination) have little effect on mental health in the 45–64 age group, which is the most productive group of older people. Also, it is important to note that the conditional probability of depression due to social discrimination goes up with age. Similarly, We found that food insecurity affects mental health the most in the 46–60 age group, which is the most productive group of older people. According to one interpretation of these findings, the impact of dissatisfaction with life on the mental health of older adults is minimal in those under 45 but steadily increases until the oldest old people. Post-75-year-old age group, again, people have minimal effect on their mental health due to a sense of dissatisfaction with life. We found that the above-mentioned connectedness factor improves mental health most for people older than 65, which are youngest-old, middle-old and older-old.

**Figure 3:**
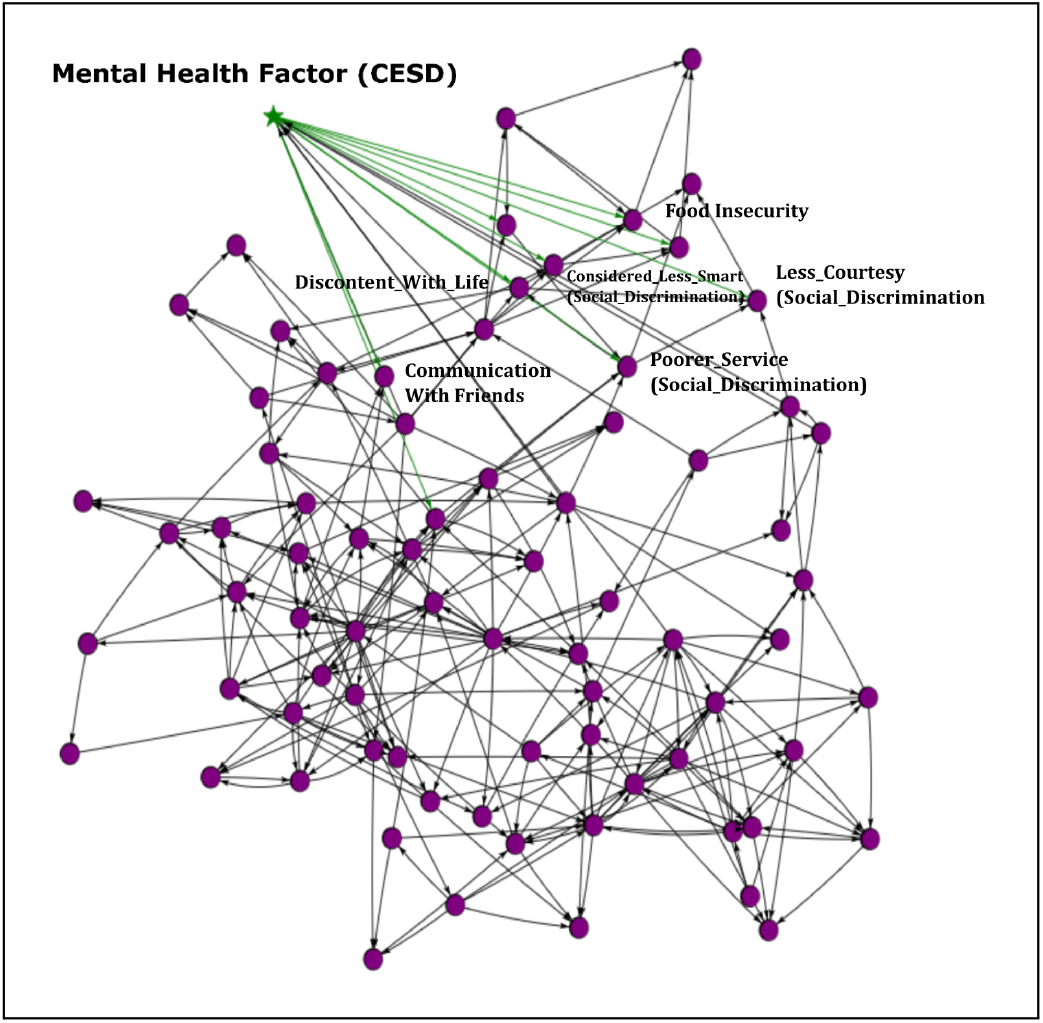
Representation of some of the association of socioeconomic factors with mental health. We learned a Bayesian Network from the LASI data, where the Starred green node represents mental health, in terms of CESD.

**Figure 4:**
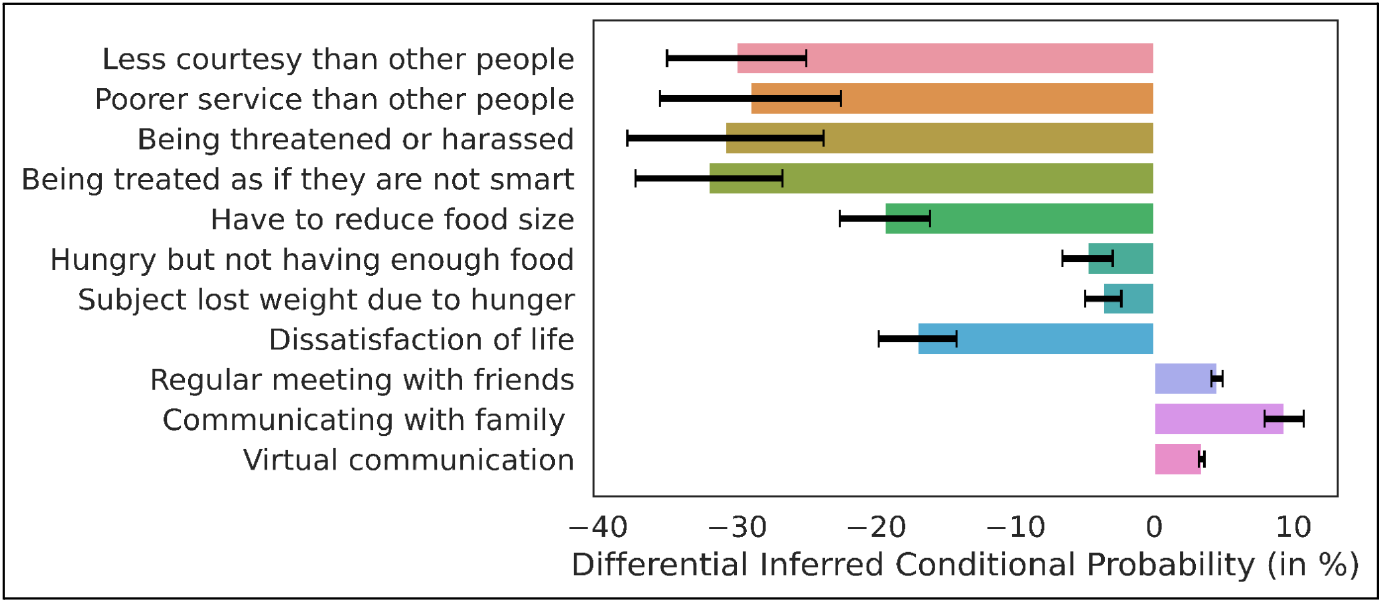
Socio-economic factors analysis with Mental illness:

**Figure 5:**
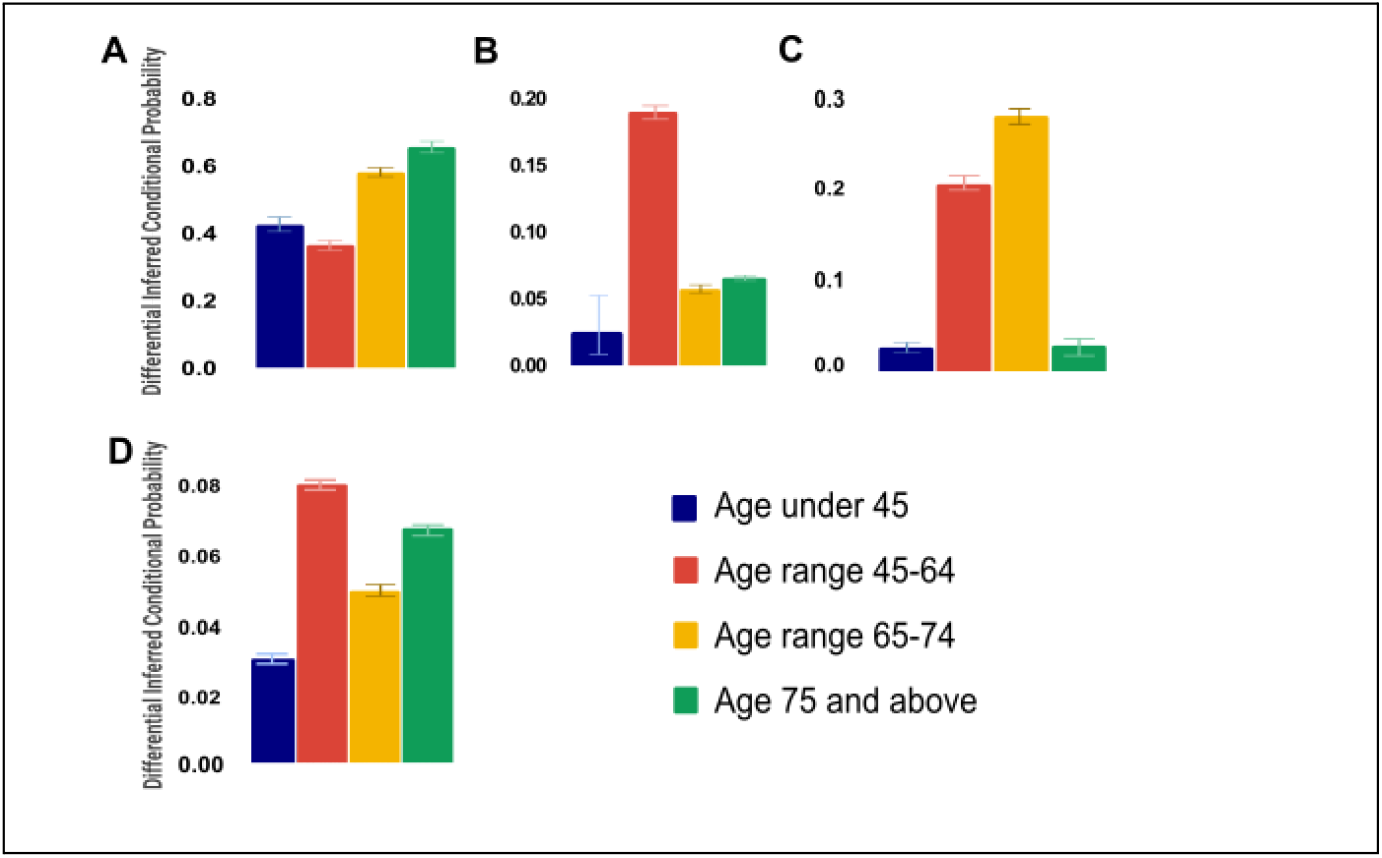
Age based stratification analysis for (A) Social Factor - Social Discrimination (B) Economical Factor - Food Insecurity (C) Psychological Factor - Discontentment about Life (D) Family and friends connectedness Factor – Communication

## Discussion

Our research explores the probabilistic graphical models that produce a list of actionable interventions and support systems for identifying the most vulnerable senior citizens in terms of mental wellbeing. The aim was to explore how common mental health issues are among older people as well as the association with various socioeconomic and psychological factors. Our analysis found that mental health is heavily linked with food insecurity, social discrimination, and psychological factors for senior citizens. Moreover, the age-based stratification analysis revealed that the multimorbidity of mental health issues increases with age, and some specific age groups are more vulnerable than others. These findings have implications for geriatric wellbeing policies in India and abroad.

The prevalence of geriatric mental health issues is a global health challenge. In this study, we revealed that food insecurity is linked to mental health issues among older people. In both developing and developed countries, there are a lot of older people who do not have enough food to eat. Over the years, the Indian government has made various efforts to reduce food insecurity and improve access to food for its citizens. Some important programs are Public Distribution System (PDS), Integrated Child Development Services Scheme (ICDS), Mid-day meal Programs (MDM), Special Nutrition Programs (SNP), Wheat Based Nutrition Programs (WNP), Applied Nutrition Programs (ANP), Antyodaya Anna Yojana (AAY). These initiatives have helped in reducing food insecurity and improving access to food for the vulnerable sections of society. However, challenges such as leakages in the PDS system and inadequate implementation of schemes remain a concern. Also, there are a few food insecurity programs available for older people in India. The Annapurna Scheme, established by the Ministry of Rural Development in 2000, targets to curb the food insecurity problem among senior citizens who are not covered by the National Old Age Pension Scheme (NOAPS) or any other pension scheme. In this programme, eligible recipients receive 10 kg of free food grains per month. Nevertheless, the desired objectives could not be attained for numerous reasons. Our study emphasizes the need for targeted interventions to address food insecurity among older people to promote better mental health outcomes.

Social discrimination is another factor linked to geriatric mental wellbeing. Our study found that older people face social discrimination in various forms, including receiving less courteous or respectful treatment, poorer service at public places like restaurants or stores, being threatened or harassed, and being treated as if they are not smart. Social discrimination can make people feel lonely, isolated, and depressed, all of which are detrimental to mental wellbeing. The Indian government has taken several initiatives to promote social inclusion and address social discrimination among older people. One such initiative is the National Policy on Senior Persons, which was formulated in 1999 and reconstituted in 2012 as the National Council of Senior Citizens. The goal was to ensure the well-being of older people and provide them with opportunities for full participation in society. The policy focuses on several areas, including health care, social security, and protection against abuse and neglect. In 2017, the government launched the Rashtriya Vayoshri Yojana (RVY), a scheme that provides assistive devices such as hearing aids, walking sticks, and spectacles to senior citizens who are below the poverty line. The aim of the scheme is to help improve the quality of life of older persons and enable them to live independently. The government has also established the National Council of Senior Citizens to advise on matters relating to the welfare of older persons and monitor the implementation of policies and programs. In addition to these initiatives, the government has taken steps to promote intergenerational bonding and reduce social isolation among older people. One such program is the Anubhav program, which encourages school students to spend time with older people and learn from their experiences. The government has also launched the Senior Citizen Savings Scheme, which provides financial security to older persons and helps them to meet their financial needs. Despite these initiatives, social discrimination and exclusion continue to be significant challenges for older people in India. There is a need for further research and policy interventions to address these issues and ensure that older people are able to live with dignity and respect in their communities. Therefore, promoting social inclusion and addressing social discrimination among older people can be an effective strategy to improve mental health outcomes.

Psychological factors such as depression, anxiety, and stress were found to be strongly associated with mental health issues among older people. Our study highlights the importance of addressing psychological factors in promoting better mental health outcomes. Psychological interventions such as cognitive-behavioral therapy and mindfulness-based stress reduction have been found to be effective in improving mental health outcomes among older people [31]. The Indian government has taken several initiatives to promote psychological well-being among older people. The National Mental Health Policy, 2014 emphasizes the need to provide mental health services to all citizens, including the elderly, and to ensure that mental health care is integrated with primary health care services. The policy also emphasizes the need for promoting mental health literacy among the elderly and their families to reduce stigma and discrimination. The National Programme for the Health Care of the Elderly (NPHCE), launched in 2010, aims to provide comprehensive health care services to elderly people, including mental health services. Under this program, special clinics for elderly patients have been set up in hospitals and health centers across the country, which provide mental health services and counseling to older people. The program also focuses on creating awareness about mental health issues among the elderly and their caregivers. The government has also launched the “National Initiative for the Care of Elderly” (NICE) program, which aims to provide a continuum of care for the elderly, including psychological and social support. The program includes the establishment of day care centers for the elderly, counseling services, and support groups for older people and their caregivers. Additionally, the government has initiated various schemes and programs to provide financial assistance to older people, including the National Old Age Pension Scheme and the Indira Gandhi National Old Age Pension Scheme, which provide monthly pension to elderly people living below the poverty line. This financial assistance can help alleviate the psychological stress associated with financial insecurity in old age. Overall, the Indian government has made significant efforts to promote psychological well-being among older people through various policies and programs. However, there is still a long way to go in terms of ensuring access to mental health services for all older people, addressing the stigma associated with mental illness, and providing comprehensive support to older people for their psychological well-being. Therefore, incorporating psychological interventions in mental health policies and practices can be an effective strategy to address mental health issues among older people.

Age-based stratification analysis revealed that multimorbidity of mental health issues increases with age. Our findings are consistent with previous studies that highlight the increasing burden of multimorbidity with age. The increasing burden of multimorbidity among older people emphasizes the need for integrated care approaches that address multiple health conditions simultaneously. Integrated care models such as the Chronic Care Model have been found to be effective in improving health outcomes among older people with multiple chronic conditions. Therefore, implementing integrated care models in mental health policies and practices can be an effective strategy to address the increasing burden of multimorbidity among older people.

Finally, it seems that mental health issues among older people are often overlooked and underdiagnosed. Our study emphasizes the need for targeted interventions to address mental health issues among older people in India. The National Program for Health Care of the Elderly (NPHCE) is an initiative by the Indian government to address the health needs of older people. The program aims to provide comprehensive health care services, including mental health care, to older people (Government of India, 2011). However, the implementation of the program faces several challenges, including inadequate funding, lack of trained health care professionals, and poor awareness among older people about mental health issues. Therefore, addressing these challenges and strengthening the NPHCE program can be an effective strategy to address mental health issues among older people in India.

Globally, mental health issues among older people are a significant public health challenge. Even in the Indian context, where the population of old individuals is projected to increase rapidly in the coming decades, our findings emphasize the urgent need for improved mental health services for the older population. In India, mental health issues among older individuals are often overlooked and undertreated, in part due to stigma surrounding mental illness [32]. Our study underscores the need for targeted interventions to improve mental health outcomes among older individuals in India, including increased access to mental health services and efforts to reduce social discrimination and food insecurity.

Overall, our study highlights the importance of addressing social determinants of health, implementing age-based stratification analysis, and developing targeted interventions to improve mental health outcomes among older individuals. Further research is needed to identify effective interventions to address the complex and multifaceted nature of mental health issues in this population.

## Data Availability

All data produced are available online at https://iipsindia.ac.in/content/LASI-data

## References

1. Luppa M, Luck T, Brähler E, König H-H, Riedel-Heller SG. Prediction of Institutionalisation in Dementia. Dement Geriatr Cogn Disord. 2008;26: 65–78. doi:10.1159/000144027

2. Weyerer S, Eifflaender-Gorfer S, Köhler L, Jessen F, Maier W, Fuchs A, et al. Prevalence and risk factors for depression in non-demented primary care attenders aged 75 years and older. J Affect Disord. 2008;111: 153–163. doi:10.1016/j.jad.2008.02.008

3. United Nations. World Population Ageing 2019 Highlights. United Nations; 2019. Available: https://play.google.com/store/books/details?id=-mz8DwAAQBAJ

4. Nations U. World population ageing 2019 highlights. U N Disarm Yearb.

5. Ruan Y, Guo Y, Zheng Y, Huang Z, Sun S, Kowal P, et al. Cardiovascular disease (CVD) and associated risk factors among older adults in six low-and middle-income countries: results from SAGE Wave 1. BMC Public Health. 2018;18: 778. doi:10.1186/s12889-018-5653-9

6. Fried LP, Ferrucci L, Darer J, Williamson JD, Anderson G. Untangling the concepts of disability, frailty, and comorbidity: implications for improved targeting and care. J Gerontol A Biol Sci Med Sci. 2004;59: 255–263. doi:10.1093/gerona/59.3.m255

7. Ferrucci L, Guralnik JM, Studenski S, Fried LP, Cutler GB Jr, Walston JD, et al. Designing randomized, controlled trials aimed at preventing or delaying functional decline and disability in frail, older persons: a consensus report. J Am Geriatr Soc. 2004;52: 625–634. doi:10.1111/j.1532-5415.2004.52174.x

8. Prince M, Bryce R, Albanese E, Wimo A, Ribeiro W, Ferri CP. The global prevalence of dementia: a systematic review and metaanalysis. Alzheimers Dement. 2013;9: 63–75.e2. doi:10.1016/j.jalz.2012.11.007

9. Skoog I. Psychiatric Disorders in the Elderly. The Canadian Journal of Psychiatry. 2011. pp. 387–397. doi:10.1177/070674371105600702

10. Skoog I. Why should general psychiatrists learn more about mental disorders in the elderly? Canadian journal of psychiatry. Revue canadienne de psychiatrie. 2011. pp. 385–386. doi:10.1177/070674371105600701

11. Waring EM. Psychiatric illness in the elderly. Am Fam Physician. 1980;21: 109–112. Available: https://www.ncbi.nlm.nih.gov/pubmed/7350731

12. Whiteford HA, Ferrari AJ, Degenhardt L, Feigin V, Vos T. Global Burden of Mental, Neurological, and Substance Use Disorders: An Analysis from the Global Burden of Disease Study 2010. In: Patel V, Chisholm D, Dua T, Laxminarayan R, Medina-Mora ME, editors. Mental, Neurological, and Substance Use Disorders: Disease Control Priorities, Third Edition (Volume 4). Washington (DC): The International Bank for Reconstruction and Development / The World Bank; doi:10.1596/978-1-4648-0426-7_ch2

13. Whiteford HA, Ferrari AJ, Degenhardt L, Feigin V, Vos T. The global burden of mental, neurological and substance use disorders: an analysis from the Global Burden of Disease Study 2010. PLoS One. 2015;10: e0116820. doi:10.1371/journal.pone.0116820

14. Global Burden of Disease Study 2013 Collaborators. Global, regional, and national incidence, prevalence, and years lived with disability for 301 acute and chronic diseases and injuries in 188 countries, 1990-2013: a systematic analysis for the Global Burden of Disease Study 2013. Lancet. 2015;386: 743–800. doi:10.1016/S0140-6736(15)60692-4

15. Stuart H. Reducing the stigma of mental illness. Glob Ment Health (Camb). 2016;3: e17. doi:10.1017/gmh.2016.11

16. Gaiha SM, Taylor Salisbury T, Koschorke M, Raman U, Petticrew M. Stigma associated with mental health problems among young people in India: a systematic review of magnitude, manifestations and recommendations. BMC Psychiatry. 2020;20: 538. doi:10.1186/s12888-020-02937-x

17. [No title]. [cited 24 Feb 2023]. Available: https://www.apa.org/pi/ses/resources/publications/age

18. Mushtaq R. Relationship Between Loneliness, Psychiatric Disorders and Physical Health? A Review on the Psychological Aspects of Loneliness. JOURNAL OF CLINICAL AND DIAGNOSTIC RESEARCH. 2014. doi:10.7860/jcdr/2014/10077.4828

19. Wu B. Social isolation and loneliness among older adults in the context of COVID-19: a global challenge. Glob Health Res Policy. 2020;5: 27. doi:10.1186/s41256-020-00154-3

20. Reiss F, Meyrose A-K, Otto C, Lampert T, Klasen F, Ravens-Sieberer U. Socioeconomic status, stressful life situations and mental health problems in children and adolescents: Results of the German BELLA cohort-study. PLoS One. 2019;14: e0213700. doi:10.1371/journal.pone.0213700

21. Warner LM, Ziegelmann JP, Schüz B, Wurm S, Tesch-Römer C, Schwarzer R. Maintaining autonomy despite multimorbidity: self-efficacy and the two faces of social support. Eur J Ageing. 2011;8: 3–12. doi:10.1007/s10433-011-0176-6

22. Ahmedani BK. Mental Health Stigma: Society, Individuals, and the Profession. J Soc Work Values Ethics. 2011;8: 41–416. Available: https://www.ncbi.nlm.nih.gov/pubmed/22211117

23. Longitudinal Ageing Study in India (LASI). [cited 24 Feb 2023]. Available: https://www.iipsindia.ac.in/lasi

24. Lee J, McGovern ME, Bloom DE, Arokiasamy P, Risbud A, O’Brien J, et al. Education, gender, and state-level disparities in the health of older Indians: Evidence from biomarker data. Econ Hum Biol. 2015;19: 145–156. doi:10.1016/j.ehb.2015.09.003

25. Website. Available: https://www.iipsindia.ac.in/lasi.

26. Radloff LS. The CES-D Scale. Applied Psychological Measurement. 1977. pp. 385–401. doi:10.1177/014662167700100306

27. Murphy KP. Machine Learning: A Probabilistic Perspective. MIT Press; 2012. Available: https://books.google.com/books/about/Machine_Learning.html?hl=&id=NZP6AQAAQBAJ

28. Koller D, Friedman N. Probabilistic Graphical Models: Principles and Techniques. MIT Press; 2009. Available: https://books.google.com/books/about/Probabilistic_Graphical_Models.html?hl=&id=7dzpHCHzNQ4C

29. Awasthi R, Patel P, Joshi V, Karkal S, Sethi T. Learning Explainable Interventions to Mitigate HIV Transmission in Sex Workers Across Five States in India. arXiv [cs.LG]. 2020. Available: http://arxiv.org/abs/2012.01930

30. Zhang M. Robust methods to improve efficiency and reduce bias in estimating survival curves in randomized clinical trials. Lifetime Data Anal. 2015;21: 119–137. doi:10.1007/s10985-014-9291-y

31. Weidman AC, Cheng JT, Tracy JL. The psychological structure of humility. Journal of Personality and Social Psychology. 2018. pp. 153–178. doi:10.1037/pspp0000112

32. Grover S, Malhotra N. Depression in elderly: A review of Indian research. Journal of Geriatric Mental Health. 2015;2: 4. doi:10.4103/2348-9995.161376

